# Deep Learning-Identified Clinical Trajectory Patterns and Associations with Kidney Outcomes in IgA Nephropathy

**DOI:** 10.1101/2025.09.04.25333884

**Authors:** Ryunosuke Noda, Daisuke Ichikawa, Sayuri Shirai, Yugo Shibagaki, Takashi Yokoo, Yusuke Suzuki, the J-IGACS Working Group

## Abstract

**Background:** The heterogeneous course of IgA nephropathy limits risk stratification based on static markers. We sought to identify clinical trajectory subgroups using unsupervised deep learning and validate their association with long-term renal outcomes.

**Methods:** We analyzed 873 biopsy-proven cases from the nationwide Japan IgA Nephropathy Prospective Cohort Study (J-IGACS). A long short-term memory autoencoder was used to generate low-dimensional representations of hematuria, proteinuria, and estimated glomerular filtration rate (eGFR) over the first 12 months after renal biopsy. We applied k-means clustering to these representations. The primary outcome was a 30% decline in eGFR from baseline.

**Results:** Three trajectory clusters were identified. Cluster 1 (n=284) showed rapid resolution of hematuria and proteinuria with stable eGFR and favorable prognosis. Cluster 2 (n=215) exhibited persistent severe hematuria, modest proteinuria reduction, and mild eGFR decline. Cluster 3 (n=374) presented with the lowest baseline eGFR and showed further decline within the first 12 months after biopsy, with incomplete proteinuria resolution despite milder hematuria. Clusters 2 and 3 had worse outcomes than Cluster 1. In Cox models adjusted for age, mean arterial pressure, and Oxford classification, cluster membership was independently associated with the primary outcome (hazard ratio 2.12; 95% CI 1.35-3.34 for Clusters 2 and 3 versus 1).

**Conclusions:** An unsupervised deep learning approach applied to trajectories of hematuria, proteinuria, and eGFR within the first year after renal biopsy identified three patient subgroups with distinct long-term renal risks. Trajectory-based classification may complement established baseline predictors and support more dynamic risk stratification in IgA nephropathy.

**Key Points:**

- Deep learning on clinical trajectories revealed the heterogeneity of IgA nephropathy, identifying three distinct patient subgroups.
- These subgroups, reflecting a spectrum of progression patterns and treatment responses, had distinct long-term renal outcomes.
- This approach may provide a dynamic framework to understand clinical course, moving beyond static, single-point risk assessment.

## Introduction

IgA nephropathy is the most common primary glomerulonephritis worldwide and has a highly heterogeneous clinical course, ranging from mild cases with spontaneous remission to severe cases progressing to end-stage kidney disease.^1,2^ This clinical diversity presents a major obstacle to accurate risk assessment and personalized medicine.^3^ However, the patterns of these heterogeneous clinical courses are not well-characterized or classified. Therefore, there is a pressing need to objectively identify distinct clinical course patterns to improve risk stratification.

Historically, predicting renal outcomes in IgA nephropathy has relied on baseline clinical and pathological markers, such as the degree of proteinuria, estimated glomerular filtration rate (eGFR), and histological findings from renal biopsy.^4–6^ However, these static, single-time-point assessments fail to capture dynamic prognostic information, such as treatment response or changes in disease activity over time. Although clinical practice is often guided by these dynamic changes, standardized approaches for classifying patients based on their evolving clinical patterns remain unestablished. To overcome this limitation, analyzing clinical marker trajectories after diagnosis has emerged as a promising approach.^7,8^

Previous studies using statistical models for trajectory analysis have revealed associations between proteinuria or hematuria trajectories and renal outcomes.^7,8^ However, these approaches often focus on a single clinical marker or analyze multiple markers individually, limiting their ability to capture the interactive dynamics among markers. Furthermore, the application of dynamic deep learning in this area has been limited. For instance, a supervised model was developed to predict renal outcomes but was not optimized for identifying objective patient subgroups.^9^

Based on these limitations, we hypothesized that distinct patient clusters could be identified from longitudinal clinical data using an unsupervised deep learning approach. To test this hypothesis, we employed a Long Short-Term Memory (LSTM) autoencoder to learn integrated trajectories of hematuria, proteinuria, and eGFR, followed by k-means clustering to identify patient subgroups. This study had two objectives: first, to identify and characterize distinct clinical trajectory clusters in a real-world setting to better understand disease heterogeneity, and second, to validate the association of these clusters with long-term renal outcomes and assess their incremental prognostic value over conventional baseline predictors.

## Methods

### Study Population

This study used data from the Japan IgA Nephropathy Prospective Cohort Study (J-IGACS), a multicenter prospective observational study conducted at 33 university hospitals and 11 regional core hospitals in Japan.^10,11^ J-IGACS is a nationwide, electronic health record-based registry of patients with biopsy-proven primary IgA nephropathy. Patients were enrolled from April 1, 2005, to August 31, 2015, and were followed up through May 31, 2021. Investigators uploaded datasets to the J-IGACS website every six months.

We included patients from the J-IGACS database who met the following criteria: baseline eGFR ≥ 30 mL/min/1.73 m²; at least 12 months of follow-up; and no more than one missing value for urinary red blood cell (RBC) count, proteinuria, or eGFR at the 0, 6, and 12-month time points used for clustering. As this was an exploratory analysis using data from the J-IGACS registry, no prior sample size calculation was performed. The study was approved by the institutional review board of St. Marianna University School of Medicine (Approval No. 6589) and the ethics committees of each participating institution. The study was conducted in accordance with the Declaration of Helsinki and adhered to the Strengthening the Reporting of Observational Studies in Epidemiology guidelines.^12^ All patients or their legal guardians provided written informed consent before enrollment.

### Variables and Outcomes

Baseline data collected at the time of renal biopsy included demographics (age, sex), clinical parameters (Body Mass Index [BMI], mean arterial pressure [MAP], eGFR, proteinuria, urinary RBC), histopathological findings (Oxford classification score), and initial treatment information (immunosuppressive therapy, tonsillectomy, Renin-Angiotensin System (RAS) inhibitor). MAP was calculated as follows: MAP (mmHg) = 1/3 × pulse pressure + diastolic blood pressure. For patients aged ≥ 20 years, eGFR was calculated using the Chronic Kidney Disease Epidemiology Collaboration (CKD-EPI) equation modified with a Japanese coefficient.^13^ For patients < 20 years old at biopsy, eGFR was calculated using the Uemura formula^14^; subsequent measurements after a patient reached 20 years of age used the modified CKD-EPI equation for Japanese adults. Urinary RBC counts were measured using either an automated urinalysis system or manual microscopic examination of the sediment, according to the standard protocol of the institution. Proteinuria was recorded from 24-hour urine collections. The Oxford classification score^15,16^ included mesangial hypercellularity (M: M0/M1), endocapillary hypercellularity (E: E0/E1), segmental glomerulosclerosis (S: S0/S1), tubular atrophy/interstitial fibrosis (T: T0/T1/T2), and cellular/fibrocellular crescents (C: C0/C1/C2), analyzed by five pathologists blinded to clinical data, with scores determined by a consensus of at least three pathologists. Initial treatment was defined as therapy initiated within one year of biopsy. Immunosuppressive therapy included oral or intravenous corticosteroids. RAS inhibitor use was defined as the initiation of an Angiotensin-Converting Enzyme inhibitor or Angiotensin II Receptor Blocker. Longitudinal data for urinary RBC, proteinuria, and eGFR at baseline, 6 months, and 12 months were used for clustering.

The primary outcome was a 30% decline in eGFR from baseline. This outcome is a widely accepted surrogate endpoint for CKD progression that provides statistical efficiency.^17–19^ Secondary outcomes for sensitivity analyses included a 40% decline in eGFR, a 50% decline in eGFR, and kidney failure (eGFR <15 mL/min/1.73 m² or initiation of kidney replacement therapy). Patients were followed from the time of renal biopsy until the loss to follow-up, or the end of the study period on May 31, 2021. Time to event was calculated in months.

### Data Preprocessing and Missing Data Handling

For model input, urinary RBC counts were converted to a five-level ordinal variable based on the thresholds commonly used in Japanese clinical practice to assess hematuria severity: 1: <5/HPF (normal/remission), 2: 5-10/HPF (mild), 3: 11-20/HPF (moderate), 4: 21-50/HPF (significant), 5: >50/HPF (severe). Missing values in the time-series data for clustering (urinary RBC, proteinuria, eGFR) were imputed using last observation carried forward or next observation carried backward if no prior value existed. Continuous variables (proteinuria, eGFR) were standardized within each fold of cross-validation using the mean and standard deviation of the training data to prevent information leakage from the test data. Missing baseline data were imputed using multivariate imputation by chained equations.

### Trajectory Pattern Identification using Deep Learning

To comprehensively capture the temporal changes of multiple clinical markers following diagnosis and initial therapeutic interventions, an LSTM autoencoder model was constructed using data from the first 12 months following renal biopsy. This deep learning architecture is particularly well-suited for capturing complex patterns in longitudinal data and has been applied for trajectory-based subgrouping in other kidney diseases, such as acute kidney injury and nephrotic syndrome.^20,21^ The model’s encoder compresses the one-year trajectories of the three biomarkers into a low-dimensional latent representation, and the decoder subsequently reconstructs the original trajectories from this representation. The ordinal urinary RBC variable was transformed into a low-dimensional vector via an Embedding layer, concatenated with standardized proteinuria and eGFR, and fed into a single-layer LSTM encoder. A detailed description of the model architecture, hyperparameter optimization process, and specific implementation is provided in the **Supplemental Methods**. Finally, the optimized model was retrained on the entire dataset to extract a latent representation for each patient’s trajectory. k-means clustering ^22^ was applied to the extracted latent representations to group patients into clusters. The optimal number of clusters (K) was determined by evaluating the silhouette score and Calinski-Harabasz index for K ranging from 2 to 6. The identified clusters were visualized in a two-dimensional space using the t-distributed Stochastic Neighbor Embedding (t-SNE) method.^23^

### Statistical Analysis

Continuous variables are presented as mean ± standard deviation (SD) or median (interquartile range [IQR]), and categorical variables as numbers (percentages). The trajectories for each cluster were visualized by plotting the bootstrapped mean (1000 resamples) and 95% confidence interval (CI) at each time point.

The association between the clusters and the primary outcome was visualized using Kaplan-Meier curves and compared using the log-rank test. To assess the independent prognostic value of the clusters, we constructed nested Cox proportional hazards models. Model 1 was a univariate model with cluster classification as the sole explanatory variable. Model 2 added key baseline clinical factors (age [per 10 years], MAP [per 10 mmHg]) as covariates. Model 3 added the Oxford classification scores to Model 2. Baseline urinary RBC, proteinuria, and eGFR values were excluded from the Cox models to avoid multicollinearity, as their trajectory information was incorporated into the cluster classification. The proportional hazards assumption was confirmed for all models using Schoenfeld residual tests.

To evaluate the incremental prognostic value of trajectory-based clusters over static predictors, we compared two nested multivariate Cox models: a “base model” including only static baseline predictors (age, MAP, eGFR, proteinuria, urinary RBC, and the Oxford classification score), and a “base + cluster model” that additionally included the cluster classification variable. Urinary RBC was included as dummy variables in both models. Model discrimination was assessed using the C-statistic (concordance index), reclassification using the category-free Net Reclassification Improvement (NRI), and discrimination improvement using the Integrated Discrimination Improvement (IDI). Differences in these metrics and their 95% CIs were calculated using bootstrapping (1000 resamples).

### Sensitivity Analysis

We conducted several sensitivity analyses to confirm the robustness of our findings. First, to assess the impact of initial treatments, Model 4 was constructed by adding immunosuppressive therapy, tonsillectomy, and RAS inhibitor to the covariates in Model 3. Second, we repeated the survival analyses for more severe renal outcomes, specifically a 40% or 50% decline in eGFR and progression to kidney failure. Third, we substituted the k-means algorithm with a Gaussian mixture model to test the stability of the clustering results. Finally, a complete case analysis, which excluded all patients with any missing baseline values, was performed to assess the impact of our data imputation methods on the Cox model results. The analyses were performed using Python (version 3.11.13), and a two-sided p-value <0.05 was considered statistically significant. Key libraries included TensorFlow, scikit-learn, Optuna, Pandas, NumPy, Lifelines, and Matplotlib. The source code is available on GitHub (URL: https://github.com/Ryunosuke1219/iga-nephropathy-lstm-autoencoder/tree/main).

## Results

### Baseline Characteristics

Of the 1,130 patients in the J-IGACS cohort, 873 met the inclusion criteria and were included in the final analysis (**Figure 1**). Exclusions were due to a baseline eGFR < 30 mL/min/1.73 m² or receipt of dialysis (n=54), a follow-up duration of < 12 months (n=92), or having two or more missing values for the clustering variables (n=111).

**Figure 1.**
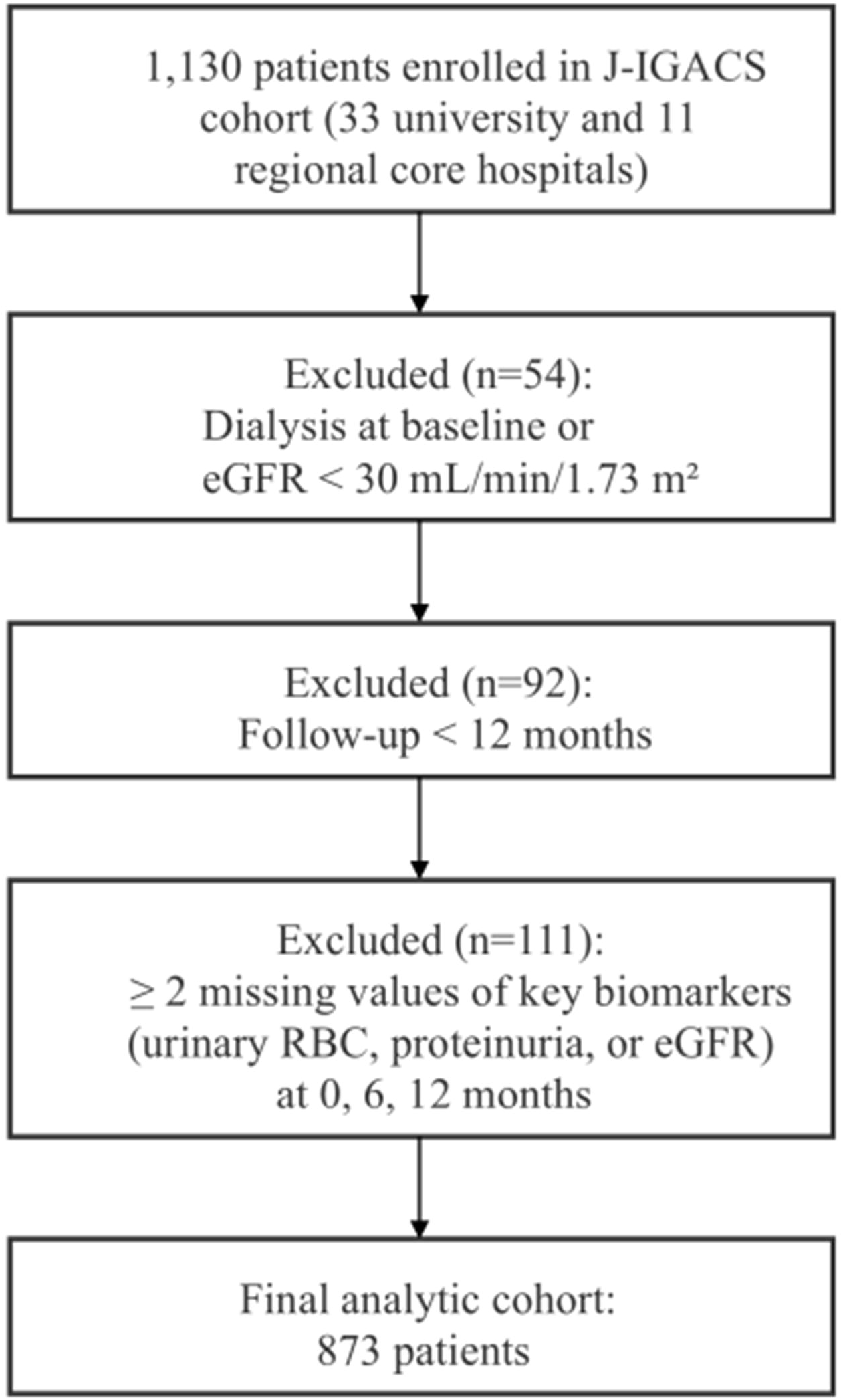
Flowchart of patient selection. Abbreviations: eGFR, estimated Glomerular Filtration Rate; J-IGACS, Japan IgA Nephropathy Prospective Cohort Study; RBC, red blood cells.

The baseline characteristics of the study population are presented in **Table 1**. The mean age was 39.0 ± 15.7 years, 50.9% of patients were female, the mean eGFR was 77.0 ± 25.9 mL/min/1.73 m², and the median proteinuria was 0.58 g/day (IQR, 0.29-1.13). Regarding the Oxford classification, 26.1% were M1, 33.6% were E1, 72.6% were S1, 18.7% were T1/T2, and 36.7% were C1/C2. For initial treatment, 65.3% received immunosuppressive therapy, 44.8% underwent tonsillectomy, and 58.3% were treated with RAS inhibitor. During a median follow-up of 78 months (IQR, 42-96), 152 patients (17.4%) experienced the primary outcome: a 30% decline in eGFR from baseline.

**Table 1.** Baseline characteristics of the study population.

| Characteristic | Overall (n=873) | Missing, n (%) |
| --- | --- | --- |
| Age (years) | 39.0 ± 15.7 | 0 (0.0) |
| Female | 444 (50.9%) | 0 (0.0) |
| Body mass index (kg/m<sup>2</sup>) | 22.2 ± 3.8 | 22 (2.5) |
| Mean arterial pressure (mmHg) | 90.0 ± 13.5 | 51 (5.8) |
| eGFR (ml/min/1.73 m<sup>2</sup>) | 77.0 ± 25.9 | 2 (0.2) |
| Proteinuria (g/day) | 0.58 [0.29-1.13] | 2 (0.2) |
| Urinary RBC (cells/HPF), n (%) |  | 10 (1.1) |
| Grade 1: <5 | 128 (14.7%) |  |
| Grade 2: 5-10 | 120 (13.7%) |  |
| Grade 3: 11-20 | 165 (18.9%) |  |
| Grade 4: 21-50 | 197 (22.6%) |  |
| Grade 5: >50 | 253 (29.0%) |  |
| M, n (%) |  | 37 (4.2) |
| M1 | 228 (26.1%) |  |
| E, n (%) |  | 37 (4.2) |
| E1 | 293 (33.6%) |  |
| S, n (%) |  | 37 (4.2) |
| S1 | 634 (72.6%) |  |
| T, n (%) |  | 37 (4.2) |
| T1 | 144 (16.5%) |  |
| T2 | 19 (2.2%) |  |
| C, n (%) |  | 37 (4.2) |
| C1 | 313 (35.9%) |  |
| C2 | 7 (0.8%) |  |
| Immunosuppressive therapy | 570 (65.3%) | 0 (0.0) |
| Tonsillectomy | 391 (44.8%) | 0 (0.0) |
| RAS inhibitor | 509 (58.3%) | 0 (0.0) |

Data are presented as mean ± standard deviation (SD), median [interquartile range (IQR)], or n (%). Abbreviations: BMI, Body Mass Index; eGFR, estimated Glomerular Filtration Rate; MAP, Mean Arterial Pressure; RAS, Renin-Angiotensin System; HPF, high-power field. The Oxford classification score consisted of mesangial hypercellularity (M0, mesangial score ≤ 0.5; M1, mesangial score > 0.5), endocapillary hypercellularity (E0, absent; E1, present), segmental glomerulosclerosis (S0, absent; S1, present), tubular atrophy/interstitial fibrosis (T0, ≤ 25%; T1, 26–50%; T2, > 50%), and crescent (C1, cellular/fibrocellular crescents present in at least one glomerulus; C2, cellular/fibrocellular crescents present in > 25% glomerulus).

### Identification of Patient Clusters Based on Clinical Trajectories

Clustering was performed on latent representations extracted by the LSTM autoencoder from the first year of longitudinal data post-biopsy. Based on the silhouette score and Calinski-Harabasz index, K=3 was determined as the optimal number of clusters (**Supplemental Figure S1**). A two-dimensional t-SNE visualization showed that the three clusters were generally well-separated (**Figure 2**). All 873 patients were classified into three clusters: Cluster 1 (n=284), Cluster 2 (n=215), and Cluster 3 (n=374).

**Figure 2.**
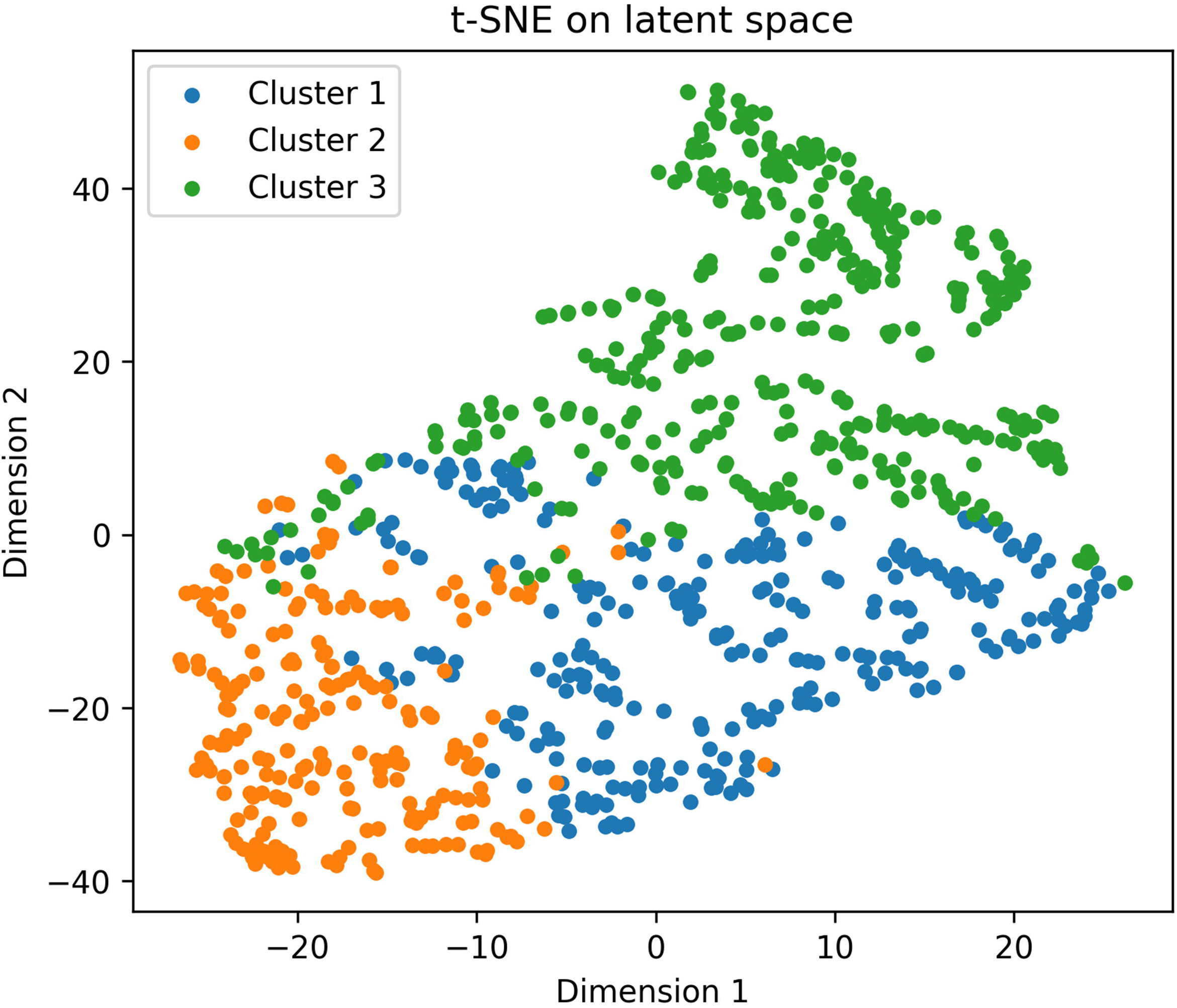
Visualization of patient clusters using t-SNE. The t-distributed Stochastic Neighbor Embedding (t-SNE) plot visualizes the low-dimensional latent representations learned by the Long Short-Term Memory (LSTM) autoencoder from the first-year clinical trajectories. Each point represents a single patient, colored according to their assigned cluster: Cluster 1 (blue), Cluster 2 (orange), and Cluster 3 (green).

### Clinical Features of the Identified Clusters

The three clusters displayed distinct clinical trajectories over the first year following renal biopsy, which were associated with baseline clinicopathological features and initial treatment choices (**Figure 3, Supplemental Table S1**).

**Figure 3.**
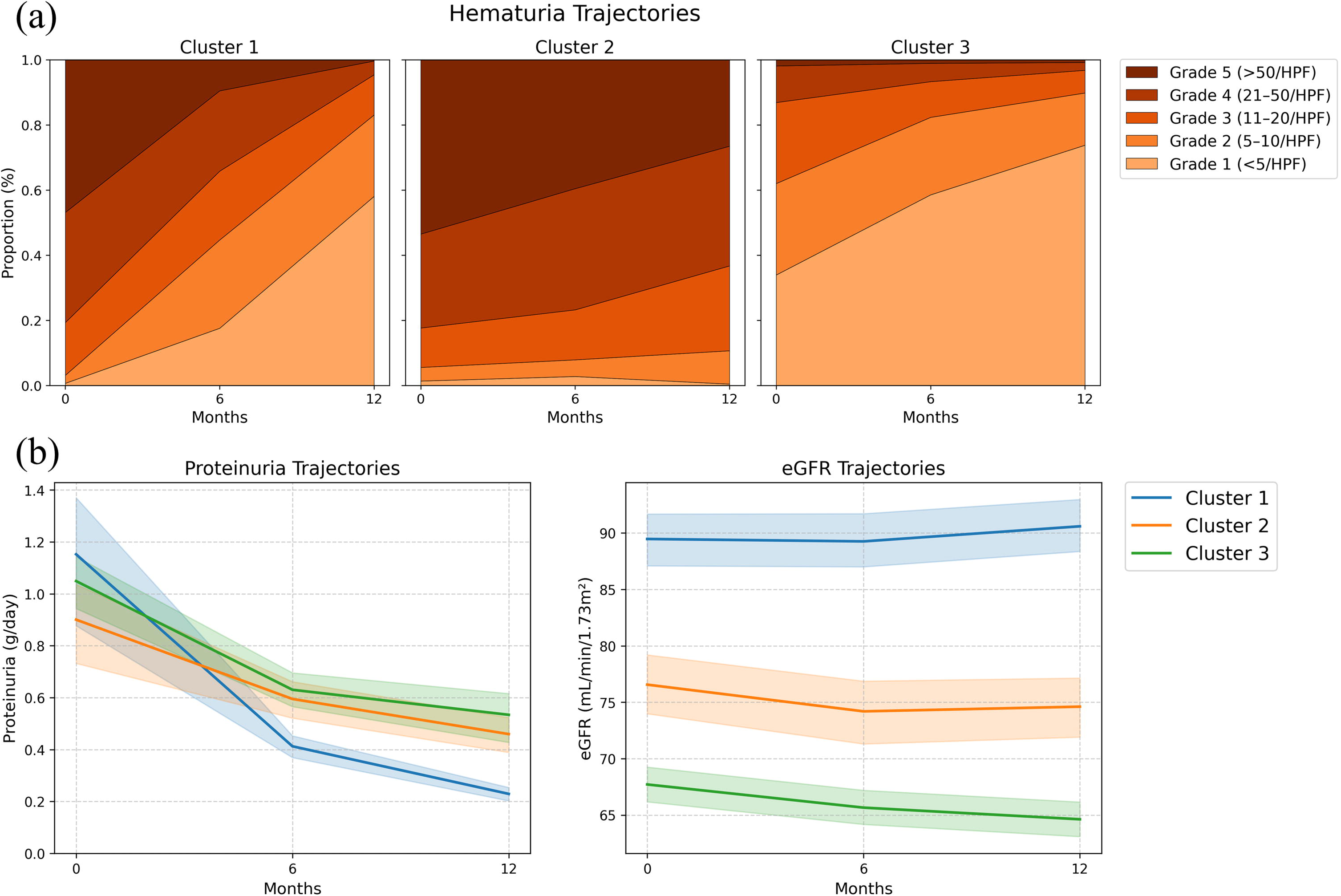
Clinical marker trajectories over the first 12 months for each cluster. (a) Hematuria trajectories are shown as stacked area plots, representing the proportion of patients within each of the five hematuria grades over 12 months for each cluster. The grades are defined by urinary red blood cells per high-power field (HPF): Grade 1 (<5/HPF), Grade 2 (5-10/HPF), Grade 3 (11-20/HPF), Grade 4 (21-50/HPF), and Grade 5 (>50/HPF). (b) Proteinuria and estimated Glomerular Filtration Rate (eGFR) trajectories are shown for each cluster. Solid lines represent the bootstrapped mean values, and the surrounding shaded areas indicate the 95% confidence intervals.

Cluster 1 was characterized by marked clinical improvement. The proportion with severe hematuria (Grade 5: > 50/HPF) decreased from 46.8% (n=133) at baseline to 0.4% (n=1) at 12 months. Similarly, mean proteinuria fell from 1.15 (95% CI, 0.88-1.37) to 0.23 g/day (95% CI, 0.20-0.25). Their eGFR, the highest at baseline, remained stable, changing negligibly from 89.5 (95% CI, 87.1-91.7) to 90.6 mL/min/1.73 m² (95% CI, 88.4-93.0).

Cluster 2 was characterized by persistent severe hematuria. This hematuria (Grade 5: > 50/HPF) persisted in a substantial portion of patients (26.5%, n=57) at 12 months. In contrast, proteinuria showed a modest reduction, with the mean decreasing from 0.90 (95% CI, 0.73-1.04) to 0.46 g/day (95% CI, 0.39-0.52). Their eGFR declined mildly from a mean of 76.6 (95% CI, 74.0-79.2) to 74.6 mL/min/1.73 m² (95% CI, 71.9-77.2).

Cluster 3 had the mildest baseline hematuria but showed the modest improvement in proteinuria, which only fell from a baseline of 1.05 (95% CI, 0.94-1.14) to 0.53 g/day (95% CI, 0.43-0.62). Their eGFR, which was already the lowest among the clusters at baseline, also declined during the first year from a mean of 67.7 (95% CI, 66.2-69.3) to 64.6 mL/min/1.73 m² (95% CI, 63.1-66.2).

These three trajectory patterns corresponded to differences in baseline characteristics (**Table 2**). Patients in Cluster 1 were younger (mean age 31.8 years), had the highest eGFR (mean 89.4 mL/min/1.73 m²), and the lowest MAP (mean 86.0 mmHg). Pathologically, this cluster had the highest proportion of crescents (C1/C2: 45.5%) and received the highest proportion of immunosuppressive therapy (81.3%) and tonsillectomy (54.6%). Patients in Cluster 2 had intermediate features regarding age (mean 40.4 years), eGFR (mean 76.6 mL/min/1.73 m²), and MAP (mean 90.4 mmHg). The proportion of immunosuppressive therapy (47.4%) and tonsillectomy (39.1%) were the lowest in this group. Patients in Cluster 3 were older (mean 43.6 years) and had the lowest eGFR at baseline (mean 67.8 mL/min/1.73 m²). Their MAP (mean 93.0 mmHg) and proteinuria (median 0.67 g/day) were the highest. Pathologically, this cluster had the highest proportion of tubular atrophy/interstitial fibrosis (T1/T2: 24.6%) and the highest proportion of RAS inhibitor (67.6%).

**Table 2.** Comparison of baseline characteristics among the identified clusters.

| Characteristic | Cluster 1 (n=284) | Cluster 2 (n=215) | Cluster 3 (n=374) |
| --- | --- | --- | --- |
| Age (years) | 31.8 ± 14.6 | 40.4 ± 16.0 | 43.6 ± 14.4 |
| Female, n (%) | 161 (56.7) | 104 (48.4) | 179 (47.9) |
| BMI (kg/m<sup>2</sup>) | 21.3 ± 4.0 | 22.5 ± 3.5 | 22.8 ± 3.6 |
| MAP (mmHg) | 86.0 ± 12.4 | 90.4 ± 14.2 | 93.0 ± 13.2 |
| eGFR (mL/min/1.73 m<sup>2</sup>) | 89.4 ± 26.5 | 76.6 ± 26.0 | 67.8 ± 21.1 |
| Proteinuria (g/day) | 0.54 [0.28-0.99] | 0.52 [0.26-1.03] | 0.67 [0.33-1.30] |
| Urinary RBC (cells/HPF), n (%) |  |  |  |
| Grade 1: <5 | 2 (0.7) | 3 (1.4) | 123 (32.9) |
| Grade 2: 5-10 | 7 (2.5) | 9 (4.2) | 104 (27.8) |
| Grade 3: 11-20 | 46 (16.2) | 26 (12.1) | 93 (24.9) |
| Grade 4: 21-50 | 95 (33.5) | 60 (27.9) | 42 (11.2) |
| Grade 5: >50 | 133 (46.8) | 113 (52.6) | 7 (1.9) |
| Oxford Classification, n (%) |  |  |  |
| M1 | 67 (23.6) | 61 (28.4) | 100 (26.7) |
| E1 | 107 (37.7) | 68 (31.6) | 118 (31.6) |
| S1 | 205 (72.2) | 149 (69.3) | 280 (74.9) |
| T1/T2 | 31 (10.9) | 40 (18.6) | 92 (24.6) |
| C1/C2 | 129 (45.5) | 71 (33.0) | 120 (32.1) |
| Initial Treatment, n (%) |  |  |  |
| Immunosuppressive therapy | 231 (81.3) | 102 (47.4) | 237 (63.4) |
| Tonsillectomy | 155 (54.6) | 84 (39.1) | 152 (40.6) |
| RAS inhibitor | 118 (41.5) | 138 (64.2) | 253 (67.6) |

Data are presented as mean ± SD, median [IQR], or n (%). Abbreviations: BMI, Body Mass Index; eGFR, estimated Glomerular Filtration Rate; MAP, Mean Arterial Pressure; RAS, Renin-Angiotensin System; RBC, Red Blood Cell; HPF, high-power field.

### Association Between Clusters and Long-Term Renal Prognosis

Kaplan-Meier analysis demonstrated that event-free survival for the primary outcome differed significantly among the three clusters (log-rank p < 0.001) (**Figure 4**). Patients in Cluster 1 had a significantly more favorable renal prognosis than those in Clusters 2 and 3.

**Figure 4.**
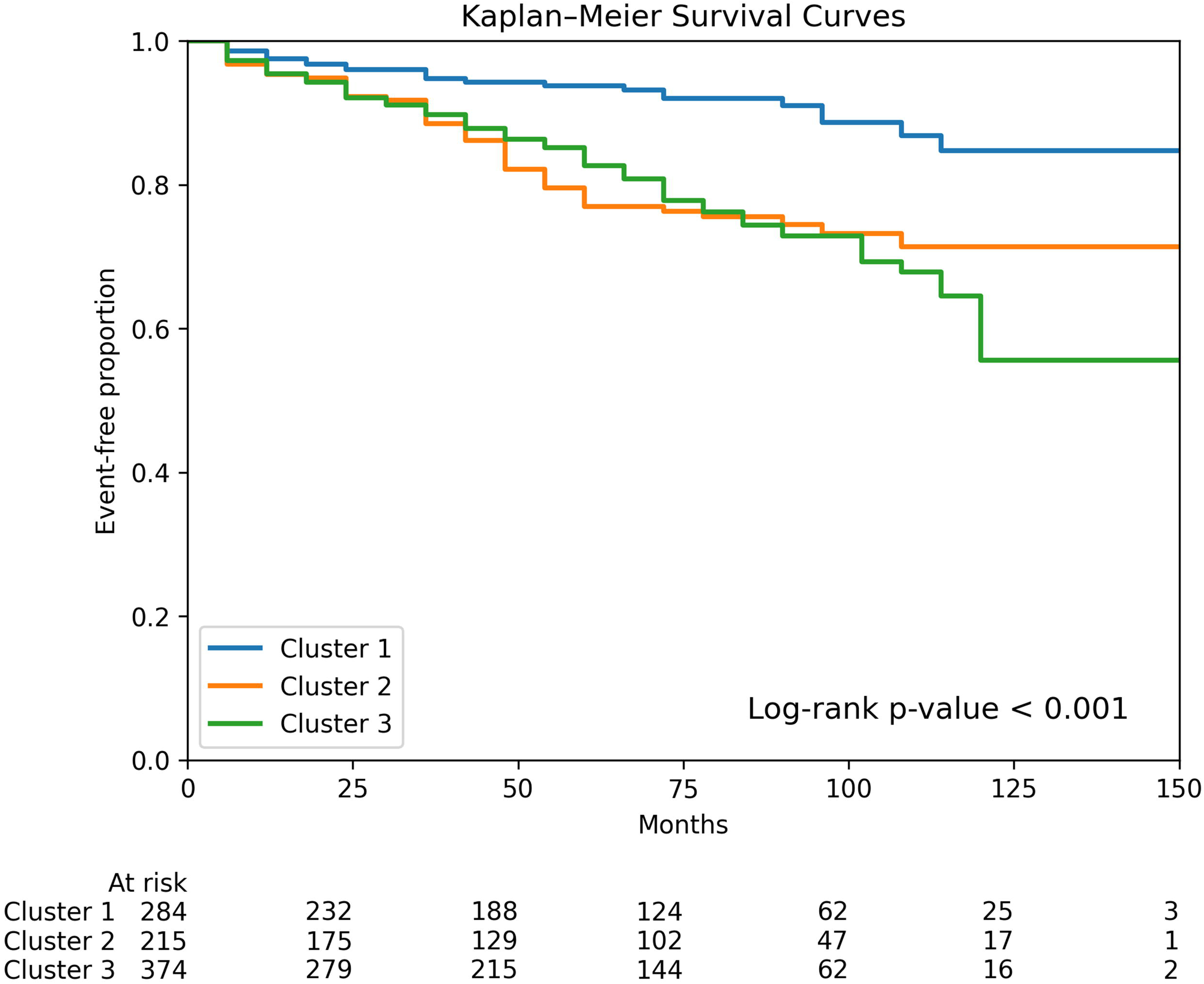
Kaplan-Meier curves for the primary outcome by cluster. Kaplan-Meier curves illustrating the event-free survival proportion for the primary outcome, defined as a ≥ 30% decline in estimated Glomerular Filtration Rate (eGFR) from baseline. The p-value was calculated using the log-rank test and showed a significant difference among the clusters (p < 0.001).

The results of the Cox proportional hazards models are presented in **Table 3**. In the unadjusted analysis (Model 1), membership in Cluster 2 or 3 was associated with a significantly higher risk for the primary outcome compared with Cluster 1 (Hazard Ratio [HR], 2.83; 95% CI, 1.83-4.37; p < 0.001). This association remained significant in Model 2, adjusted for age and MAP (adjusted HR, 2.16; 95% CI, 1.38-3.40; p < 0.001), and in Model 3, further adjusted for the Oxford classification score (adjusted HR, 2.12; 95% CI, 1.35-3.34; p = 0.001).

**Table 3.**
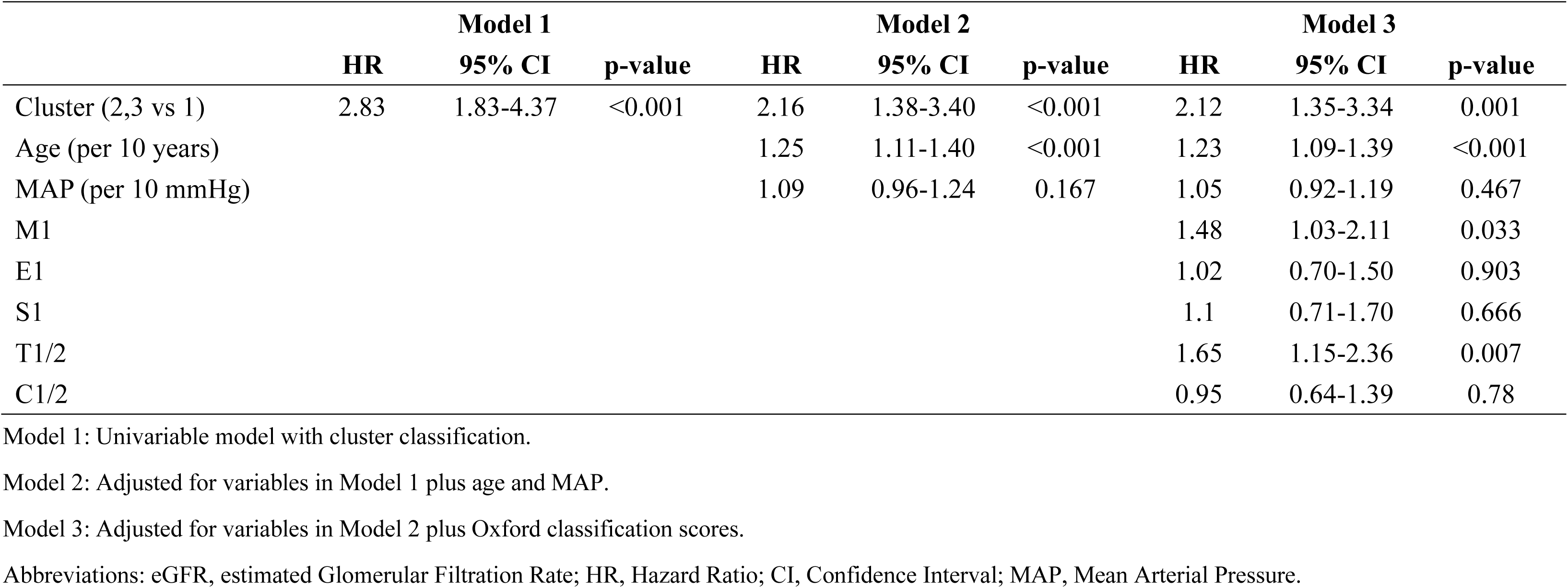
Cox proportional hazards models for the primary outcome (30% decline in eGFR)

To evaluate the incremental prognostic value of the trajectory-based classification over established static predictors, we compared nested Cox models (**Table 4**). Adding the cluster classification to the base model significantly improved model discrimination. The C-statistic increased from 0.745 (95% CI, 0.707-0.765) for the base model to 0.757 (95% CI, 0.722-0.776) for the base + cluster model (Δ, 0.013; 95% CI, 0.003-0.022). A significant improvement was also observed for the IDI (absolute value, 0.142; 95% CI, 0.028-0.307). However, the category-free NRI was not statistically significant (0.002; 95% CI, −0.026 to 0.262).

**Table 4.** Improvement in model performance for predicting 30% eGFR decline after adding cluster classification.

|  | **C statistic**<br>**(95%CI)** | **Change in C**<br>**statistic (95%CI)** | **Category-free NRI**<br>**(95%CI)** | **Absolute IDI**<br>**(95%CI)** | **Relative IDI**<br>**(95%CI)** |
| --- | --- | --- | --- | --- | --- |
| Base model | 0.745 (0.707-0.765) |  |  |  |  |
| Base + Cluster<br>model | 0.757 (0.722-0.776) | 0.013 (0.003-0.022) | 0.002 (-0.026-0.262) | 0.142 (0.028-0.307) | 0.151 (0.028-0.347) |

The Base model included static baseline factors (age, MAP, eGFR, proteinuria, urinary RBC, and the Oxford classification score). The Base +Cluster model added the cluster classification (Clusters 2 and 3 vs Cluster 1) to the base model. Abbreviations: CI, Confidence Interval; NRI, Net Reclassification Improvement; IDI, Integrated Discrimination Improvement.

### Sensitivity Analysis

The independent prognostic value of the clusters was maintained after adjusting for initial treatments (immunosuppressive therapy, tonsillectomy, and RAS inhibitors) (Model 4; adjusted HR, 1.90; 95% CI, 1.19-3.02) (**Supplemental Table S2**). Furthermore, significant prognostic separation among clusters was observed for more severe renal outcomes—40% eGFR decline, 50% eGFR decline, and kidney failure (log-rank test p < 0.05 for all) (**Supplemental Figures S2-S4**). Using a Gaussian mixture model instead of k-means also yielded prognostically significant clusters (p < 0.001) (**Supplemental Figure S5**). Finally, a complete case analysis yielded consistent results in the Cox model, confirming the independent prognostic ability of the clusters (**Supplemental Table S3**).

## Discussion

In this large, real-world cohort study, we applied an unsupervised deep learning approach to one-year trajectories of hematuria, proteinuria, and eGFR in patients with IgA nephropathy. This method identified three distinct patient clusters that were independently associated with long-term renal outcomes. These findings, which reveal subgroups with different clinical dynamics and risks, challenge a uniform management approach and underscore the need for more personalized therapeutic strategies.

The identified clusters likely represent distinct clinicopathological phenotypes within IgA nephropathy. Cluster 1, for example, appears to be a subgroup with highly active, treatment-responsive inflammation. This cluster was characterized by severe hematuria and a high prevalence of crescents, coupled with the highest rates of immunosuppressive therapy and tonsillectomy. This combination may have led to the improvement in hematuria and proteinuria and, consequently, a favorable long-term renal outcome. Prior studies have shown that crescents were a marker of severe, active glomerular inflammation and are associated with a high risk of progression.^24,25^ Our findings suggest that this high-risk state is not fixed; instead, the prognosis can be modified by effective therapeutic intervention. This highlights the importance of the interplay between the disease’s biology and treatment responsiveness, which cannot be captured by static predictors alone.

In contrast, Cluster 3 likely represents a patient group in whom established, irreversible chronic changes are the primary driver of progression. This cluster included the oldest age, the lowest baseline eGFR, and the highest proportion of T-lesions, a robust predictor of progression.^16,26^ The poor renal outcome in this cluster, despite having the highest rate of RAS inhibitor use, suggests a treatment-resistant pathophysiology for which current standard-of-care is likely insufficient. This aligns with the concept of renal fibrosis as the final common pathway of CKD progression.^27^ Therefore, patients with this trajectory may be ideal candidates for novel therapeutic agents, including anti-fibrotic therapies.

Cluster 2, characterized by persistent hematuria, is also a clinically compelling subgroup. While the prognostic value of hematuria has been debated, recent studies suggest that persistent or time-averaged microscopic hematuria is an independent risk factor for progression.^8,28–30^ The trajectory of Cluster 2 may therefore indicate a potential blind spot in current guidelines, which primarily focus on proteinuria as the main therapeutic target.^4^ Patients in this cluster whose proteinuria might fall below therapeutic thresholds could be at risk of being undertreated, representing a potential unmet medical need.

Our approach builds upon previous work. While prior clustering studies in IgA nephropathy have provided valuable insights, they have relied on static, cross-sectional data.^31,32^ Previous trajectory analyses in IgA nephropathy, however, have often focused on single markers or used statistical models that assume predefined trajectory shapes, which may not fully capture clinical complexity.^7,8^ Our study addresses these limitations by employing an LSTM autoencoder, a deep learning architecture specifically designed to capture complex patterns from longitudinal data without such rigid assumptions.^33,34^ While standard LSTM networks have recently shown efficacy in predicting outcomes in IgA nephropathy^9^, the use of an LSTM autoencoder for data-driven subgrouping represents a novel application in this field. To the best of our knowledge, this is the first study to leverage this approach to classify the diverse clinical trajectories of the disease, offering a new framework for patient stratification. This dynamic classification could move beyond static risk factors to identify clinically distinct phenotypes, such as treatment-resistant progressors or those with persistent inflammation, thereby enabling a more personalized approach to patient monitoring and therapeutic strategies.

This study has several limitations. First, as an observational study, it is susceptible to confounding by indication. The identified clusters could reflect a combination of disease biology and real-world treatment choices, not pure natural history. Accordingly, the main utility of our approach may be less in improving risk prediction and more in providing a framework to understand the diversity of clinical course. Second, this study was limited to a Japanese cohort where both patient backgrounds and clinical management may differ from other regions. Therefore, while our sensitivity analysis confirmed that the identified patterns are methodologically robust to the choice of clustering algorithm, the generalizability of these specific clinical trajectories requires validation in diverse, international cohorts. Third, our analysis was limited to three clinical markers and did not include the trajectories of other potentially important variables, such as blood pressure or specific medication dosages. These factors, combined with the inherent black-box nature of deep learning models and the limited temporal resolution of the data, imply that the precise factors driving the clustering remain to be fully elucidated. Future work should focus on validating these patterns in diverse, international cohorts and refining the model with additional clinical variables.

In conclusion, this study demonstrated that an unsupervised deep learning approach can identify clinically meaningful patient clusters in IgA nephropathy based on their longitudinal clinical data. This data-driven classification represents a critical step toward a more dynamic and personalized risk assessment. Ultimately, this approach could enable clinicians to better tailor monitoring and therapeutic strategies to a patient’s evolving clinical course, thereby advancing the goal of precision medicine in nephrology.

## Data Availability

The data underlying this article will be shared upon reasonable request to the corresponding author. The source code used for the analysis is publicly available on GitHub at　https://github.com/Ryunosuke1219/iga-nephropathy-lstm-autoencoder/tree/main.

## Disclosures

Yusuke Suzuki has received consulting fees from Otsuka Pharmaceutical (Visterra), Novartis, Chinook Therapeutics, ARGENX, BioCryst, Alexion Pharmaceuticals, Renalys, Alpine, and George Clinical.

Yusuke Suzuki has also received honoraria for lectures, presentations, manuscript writing, or educational events from Kyowa Kirin, Novartis, Mitsubishi Tanabe, Otsuka Pharmaceutical, Daiichi Sankyo, AstraZeneca, Boehringer Ingelheim, and Chinook Therapeutics.

## Funding

This study was partly supported by a Grant-in-Aid for Progressive Renal Diseases Research, Research on Rare and Intractable Disease, from the Ministry of Health, Laboure and Welfare of Japan. This research was supported by the Japan Agency for Medical Research and Development under grant JP19ek0109261.

## Acknowledgments

We thank the clinicians in the J-IGACS Working Group for their kind cooperation. The J-IGACS Working Group: Yusuke Suzuki: Department of Nephrology, Juntendo University Faculty of Medicine, Tokyo, Japan. Takashi Yokoo: Division of Nephrology and Hypertension, Department of Internal Medicine, The Jikei University School of Medicine, Tokyo, Japan. Ryosuke Aoki: Department of Nephrology, Juntendo University Faculty of Medicine, Tokyo, Japan. Daisuke Ichikawa: Division of Nephrology and Hypertension, Department of Internal Medicine, St. Marianna University School of Medicine, Kanagawa, Japan. Takafumi Ito: Department of Internal Medicine, Nephrology, Teikyo University School of Medicine, Teikyo University Chiba Medical Center, Chiba, Japan. Hiroyuki Ueda: Division of Nephrology and Hypertension, Department of Internal Medicine, The Jikei University School of Medicine, Tokyo, Japan. Maki Urushihara: Department of Pediatrics, Institute of Biomedical Sciences, Tokushima University Graduate School, Tokushima, Japan. Ritsuko Katafuchi: Kidney Unit, National Hospital Organization, Fukuoka-Higashi Medical Center, Fukuoka, Japan, and Division of Nephrology, Medical Corporation Houshikai, Kano Hospital, Fukuoka, Japan. Tetsuya Kawamura: Division of Nephrology and Hypertension, Department of Internal Medicine, The Jikei University School of Medicine, Tokyo, Japan. Masao Kikuchi: Division of Cardiovascular Medicine and Nephrology, Department of Internal Medicine, Faculty of Medicine, University of Miyazaki, Miyazaki, Japan. Masao Kihara: Department of Nephrology, Juntendo University Faculty of Medicine, Tokyo, Japan. Kentaro Koike: Division of Nephrology and Hypertension, Department of Internal Medicine, The Jikei University School of Medicine, Tokyo, Japan. Ryoko Sakaguchi: Department of Pathology, The Jikei University School of Medicine, Tokyo, Japan. Takaya Sasaki: Division of Nephrology and Hypertension, Department of Internal Medicine, The Jikei University School of Medicine, Tokyo, Japan. Satoru Sanada: Department of Nephrology, Japan Community Healthcare Organization Sendai Hospital, Sendai, Japan. Takanori Shibata: Division of Nephrology, Department of Medicine, Showa Medical University School of Medicine, Tokyo, Japan. Yuko Shima: Department of Pediatrics, Wakayama Medical University, Wakayama City, Wakayama, Japan. Akihiro Shimizu: Division of Nephrology and Hypertension, Department of Internal Medicine, The Jikei University School of Medicine, Tokyo, Japan. Akira Shimizu: Department of Analytic Human Pathology, Nippon Medical School, Tokyo, Japan. Kensuke Joh: Department of Pathology, The Jikei University School of Medicine, Tokyo, Japan. Sayuri Shirai: Division of Nephrology and Hypertension, Department of Internal Medicine, St. Marianna University School of Medicine, Yokohama City Seibu Hospital, Yokohama, Japan. Hitoshi Suzuki: Department of Nephrology, Juntendo University Faculty of Medicine, Tokyo, Japan. Kazuo Takahashi: Department of Biomedical Molecular Sciences, School of Medicine, Fujita Health University, Nagoya, Aichi, Japan. Nobuo Tsuboi: Division of Nephrology and Hypertension, Department of Internal Medicine, The Jikei University School of Medicine, Tokyo, Japan. Yasuhiko Tomino: Asian Pacific Renal Research Promotion Office, Medical Corporation SHOWAKAI, Shinjuku-ku, Tokyo, Japan. Shinya Nakatani: Department of Metabolism, Endocrinology and Molecular Medicine, Osaka Metropolitan University Graduate School of Medicine, Osaka, Japan. Koichi Nakanishi: Department of Child Health and Welfare (Pediatrics), Graduate School of Medicine, University of the Ryukyus, Ginowan, Okinawa, Japan. Masako Nishikawa: Center for Research Promotion, The Jikei University School of Medicine, Tokyo, Japan. Tomoya Nishino: Department of Nephrology, Graduate School of Biomedical Sciences, Nagasaki University, Nagasaki, Japan.

Yoshihito Nihei: Department of Nephrology, Juntendo University Faculty of Medicine, Tokyo, Japan. Akinori Hashiguchi: Department of Pathology, Keio University School of Medicine, Tokyo, Japan. Hiroshi Hataya: Department of Nephrology and Rheumatology, Tokyo Metropolitan Children’s Medical Center, Fuchu, Tokyo, Japan. Keita Hirano: Division of Nephrology and Hypertension, Department of Internal Medicine, The Jikei University School of Medicine, Tokyo, Japan. Yusuke Fukao: Department of Nephrology, Juntendo University Faculty of Medicine, Tokyo, Japan. Akihiro Fukuda: Department of Endocrinology, Metabolism, Rheumatology and Nephrology, Faculty of Medicine, Oita University, Oita, Japan. Shouichi Fujimoto: Department of Medical Environment Innovation, Faculty of Medicine, University of Miyazaki, Miyazaki, Japan. Shiko Honma: Department of Pathology, The Jikei University School of Medicine, Tokyo, Japan. Keiichi Matsuzaki: Department of Public Health, Kitasato University School of Medicine, Kanagawa, Japan. Kenichiro Miura: Department of Pediatric Nephrology, Tokyo Women’s Medical University, Tokyo, Japan. Yoichi Miyazaki: Division of Nephrology and Hypertension, Department of Internal Medicine, The Jikei University School of Medicine, Tokyo, Japan. Kumiko Muta: Advanced Medical Education Center, Nagasaki University School of Medicine, Nagasaki, Japan. Takahito Moriyama: Department of Nephrology, Tokyo Medical University, Tokyo, Japan. Takashi Yasuda: Naruse Kidney Clinic, Tokyo, Japan. Yoshinari Yasuda: Department of Advanced Science in Renal-Cardio Medicine/Nephrology, Gifu University Graduate School of Medicine, Gifu, Japan. Shinya Yokote: Department of Nephrology, Kawaguchi Municipal Medical Center, Saitama, Japan.

## Author Contributions

Conceptualization: Ryunosuke Noda, Daisuke Ichikawa.

Data curation: Ryunosuke Noda, Daisuke Ichikawa, Sayuri Shirai.

Formal analysis: Ryunosuke Noda.

Funding acquisition: Yusuke Suzuki.

Investigation: Ryunosuke Noda.

Methodology: Ryunosuke Noda.

Supervision: Daisuke Ichikawa, Sayuri Shirai, Yugo Shibagaki.

Writing – original draft: Ryunosuke Noda, Daisuke Ichikawa, Sayuri Shirai.

Writing – review & editing: Ryunosuke Noda, Daisuke Ichikawa, Sayuri Shirai, Yugo Shibagaki, Takashi Yokoo, Yusuke Suzuki.

## Notes

### Author Declarations

The study was approved by the institutional review board of St. Marianna University School of Medicine (Approval No. 6589) and the ethics committees of each participating institution. The study was conducted in accordance with the Declaration of Helsinki and adhered to the Strengthening the Reporting of Observational Studies in Epidemiology guidelines.

### Summary of Updates

We have revised the title to make the study's content clearer to the readers.

