## Supplementary Material for "Deep Learning-Identified Clinical Trajectory Patterns and Associations with Kidney Outcomes in IgA Nephropathy"

**Table of Contents**

Supplemental Methods
Table S1: Temporal changes in clinical markers for each cluster
Table S2: Cox regression analysis including initial treatments as covariates
Table S3: Cox regression analysis on a complete case basis
Figure S1: Validation of the optimal number of clusters
Figure S2: Kaplan-Meier curves for a 40% decline in eGFR
Figure S3: Kaplan-Meier curves for a 50% decline in eGFR
Figure S4: Kaplan-Meier curves for kidney failure
Figure S5: Clustering analysis by Gaussian Mixture Model
STROBE Checklist

### Supplemental Methods

#### Feature Extraction using a Long Short-Term Memory (LSTM) Autoencoder

To extract the essential features from each patient's clinical course, we used a model known as an LSTM autoencoder. This model integrates two deep learning concepts: Long Short-Term Memory (LSTM) networks and autoencoders, making it particularly suitable for analyzing longitudinal data.[1,2]

Long Short-Term Memory (LSTM) is a type of recurrent neural network adept at learning from sequential data, such as clinical measurements collected over several months. Unlike standard neural networks, LSTMs have internal memory mechanisms that allow them to recognize and remember patterns over long durations, making them ideal for capturing the evolving trajectory of a patient's condition.[3]

An autoencoder is an unsupervised neural network designed for dimensionality reduction and feature learning. It consists of two components: an encoder that compresses high-dimensional input data into a low-dimensional, information-rich summary called a latent representation, and a decoder that attempts to reconstruct the original data from this compressed representation. Through this process of compression and reconstruction, the model is forced to learn the most salient features of the data, which are captured in the latent representation.[4]

The architecture of our LSTM autoencoder began with an input tensor of shape (873, 3, 3), representing data for 873 patients with three features (urinary red blood cell count, proteinuria, and eGFR) at three time points (0, 6, and 12 months). The quantitative variables, proteinuria and eGFR, were standardized to have a mean of 0 and a standard deviation of 1. To prevent data leakage, this scaling was performed within each fold of our cross-validation scheme, where the scaler was trained on the training data only and subsequently applied to the validation set.

The encoder first processed the ordinal urinary RBC variable through an Embedding layer to transform it into a low-dimensional dense vector. This embedding vector was then concatenated with the two standardized quantitative variables. The resulting combined time-series data was passed into a single LSTM layer to encode sequential context. Finally, the output of this LSTM layer was compressed into a single latent representation vector via a Dense layer with a linear activation function.

For the decoder, the latent representation vector was duplicated by a RepeatVector layer to match the original input sequence length. This repeated vector sequence was then fed into another LSTM layer, structured identically to the encoder's, to reconstruct the temporal information. A final TimeDistributed (Dense) layer was responsible for reconstructing the original three features at each time point.

#### Hyperparameter Optimization

To maximize model performance, we systematically tuned the key hyperparameters of the LSTM autoencoder using the Optuna library, a Bayesian optimization framework. The objective of this optimization was to minimize the mean squared reconstruction error (MSE) on the validation sets across a 5-fold cross-validation scheme. The search space for the hyperparameters included an embedding layer output dimension ranging from 2 to 4, a number of LSTM units selected from {16, 32, 64}, a latent space dimension between 3 and 12, and a dropout rate for the LSTM layer from 0.05 to 0.25. During each optimization trial, the model was trained for up to 200 epochs using the Adam optimizer with a learning rate of 1e-3. We applied an Early Stopping callback to halt training if the validation loss did not improve for 15 consecutive epochs, ensuring that the best model weights were saved. The final autoencoder model was then retrained on the entire dataset using the best hyperparameter set identified through this process to extract the latent representation for each patient.

**Clustering of Clinical Trajectories and Determination of the Optimal Number of Clusters**

The extracted latent representations were subjected to K-means, a non-hierarchical clustering algorithm, to partition patients into subgroups based on the similarity of their clinical trajectory patterns.[5] The K-means algorithm operates iteratively by first initializing a pre-specified number (k) of cluster centroids at random. Next, each data point is assigned to its nearest centroid. The centroids are then recalculated as the mean of all points within each newly formed cluster. This process of assignment and recalculation is repeated until the cluster memberships stabilize. The optimal number of clusters (k) for this study was determined by systematically evaluating the silhouette score and the Calinski-Harabasz index for k ranging from 2 to 6.

**Table S1. Temporal changes in clinical markers for each cluster**

| Marker | Cluster | 0 months | 6 months | 12 months |
|---|---|---|---|---|
| eGFR (mL/min/1.73 m²) | Cluster 1 | 89.5 (87.1-91.7) | 89.3 (87.0-91.7) | 90.6 (88.4-93.0) |
| | Cluster 2 | 76.6 (74.0-79.2) | 74.2 (71.3-76.9) | 74.6 (71.9-77.2) |
| | Cluster 3 | 67.7 (66.2-69.3) | 65.7 (64.2-67.2) | 64.6 (63.1-66.2) |
| Proteinuria (g/day) | Cluster 1 | 1.15 (0.88-1.37) | 0.41 (0.37-0.45) | 0.23 (0.20-0.25) |
| | Cluster 2 | 0.90 (0.73-1.04) | 0.60 (0.52-0.66) | 0.46 (0.39-0.52) |
| | Cluster 3 | 1.05 (0.94-1.14) | 0.63 (0.57-0.70) | 0.53 (0.43-0.62) |
| Urinary RBC Grade 1 (<5 cells/HPF) | Cluster 1 | 2 (0.7) | 50 (17.6) | 165 (58.1) |
| | Cluster 2 | 3 (1.4) | 6 (2.8) | 1 (0.5) |
| | Cluster 3 | 127 (34.0) | 219 (58.6) | 276 (73.8) |
| Urinary RBC Grade 2 (5 -10 cells/HPF) | Cluster 1 | 7 (2.5) | 77 (27.1) | 71 (25.0) |
| | Cluster 2 | 9 (4.2) | 11 (5.1) | 22 (10.2) |
| | Cluster 3 | 105 (28.1) | 89 (23.8) | 60 (16.0) |
| Urinary RBC Grade 3 (11 - 20 cells/HPF) | Cluster 1 | 46 (16.2) | 60 (21.1) | 35 (12.3) |
| | Cluster 2 | 26 (12.1) | 33 (15.3) | 56 (26.0) |
| | Cluster 3 | 93 (24.9) | 41 (11.0) | 26 (7.0) |
| Urinary RBC Grade 4 (21 - 50 cells/HPF) | Cluster 1 | 96 (33.8) | 70 (24.6) | 12 (4.2) |
| | Cluster 2 | 62 (28.8) | 80 (37.2) | 79 (36.7) |
| | Cluster 3 | 42 (11.2) | 21 (5.6) | 9 (2.4) |
| Urinary RBC Grade 5 (>50 cells/HPF) | Cluster 1 | 133 (46.8) | 27 (9.5) | 1 (0.4) |
| | Cluster 2 | 115 (53.5) | 85 (39.5) | 57 (26.5) |
| | Cluster 3 | 7 (1.9) | 4 (1.1) | 3 (0.8) |

Data for eGFR and proteinuria are presented as mean (95% Confidence Interval). Data for urinary RBC grades are presented as n (%). Abbreviations: eGFR, estimated Glomerular Filtration Rate; RBC, Red Blood Cell; HPF, high-power field.

**Table S2. Cox regression analysis including initial treatments as covariates**

| | Model 4 | | |
|---|---|---|---|
| | **HR** | **95% CI** | **p-value** |
| Cluster (2,3 vs 1) | 1.90 | 1.19-3.02 | 0.007 |
| Age (per 10 years) | 1.17 | 1.03-1.32 | 0.015 |
| MAP (per 10 mmHg) | 1.01 | 0.88-1.15 | 0.942 |
| M1 | 1.39 | 0.97-1.99 | 0.076 |
| E1 | 1.04 | 0.71-1.53 | 0.841 |
| S1 | 1.14 | 0.73-1.77 | 0.563 |
| T1/2 | 1.64 | 1.14-2.36 | 0.007 |
| C1/2 | 1.01 | 0.68-1.49 | 0.955 |
| Immunosuppressive therapy | 0.86 | 0.59-1.27 | 0.449 |
| Tonsillectomy | 0.73 | 0.49-1.08 | 0.117 |
| RAS inhibitor | 1.55 | 1.02-2.36 | 0.041 |

Model 4 was adjusted for all variables in Model 3 (age, MAP, Oxford score) plus initial treatments (immunosuppressive therapy, tonsillectomy, and RAS inhibitor use).
Abbreviations: HR, Hazard Ratio; CI, Confidence Interval; MAP, Mean Arterial Pressure; RAS, Renin-Angiotensin System.

**Table S3. Cox regression analysis on a complete case basis**

| | Model 1 | | | Model 2 | | | Model 3 | | |
|---|---|---|---|---|---|---|---|---|---|
| | **HR** | **95% CI** | **p-value** | **HR** | **95% CI** | **p-value** | **HR** | **95% CI** | **p-value** |
| Cluster (2,3 vs 1) | 2.83 | 1.83-4.37 | <0.001 | 2.17 | 1.37-3.44 | 0.001 | 2.22 | 1.38-3.56 | <0.001 |
| Age (per 10 years) | | | | 1.24 | 1.10-1.39 | <0.001 | 1.2 | 1.06-1.35 | 0.003 |
| MAP (per 10 mmHg) | | | | 1.09 | 0.97-1.24 | 0.159 | 1.07 | 0.94-1.21 | 0.321 |
| M1 | | | | | | | 1.44 | 1.00-2.08 | 0.049 |
| E1 | | | | | | | 1.04 | 0.70-1.54 | 0.838 |
| S1 | | | | | | | 1.06 | 0.68-1.65 | 0.805 |
| T1/2 | | | | | | | 1.7 | 1.17-2.47 | 0.005 |
| C1/2 | | | | | | | 0.94 | 0.64-1.39 | 0.761 |

This sensitivity analysis excluded all patients with any missing baseline values. Model 1: Univariable model with cluster classification. Model 2: Adjusted for variables in Model 1 plus age and MAP. Model 3: Adjusted for variables in Model 2 plus Oxford classification scores. Abbreviations: HR, Hazard Ratio; CI, Confidence Interval; MAP, Mean Arterial Pressure.

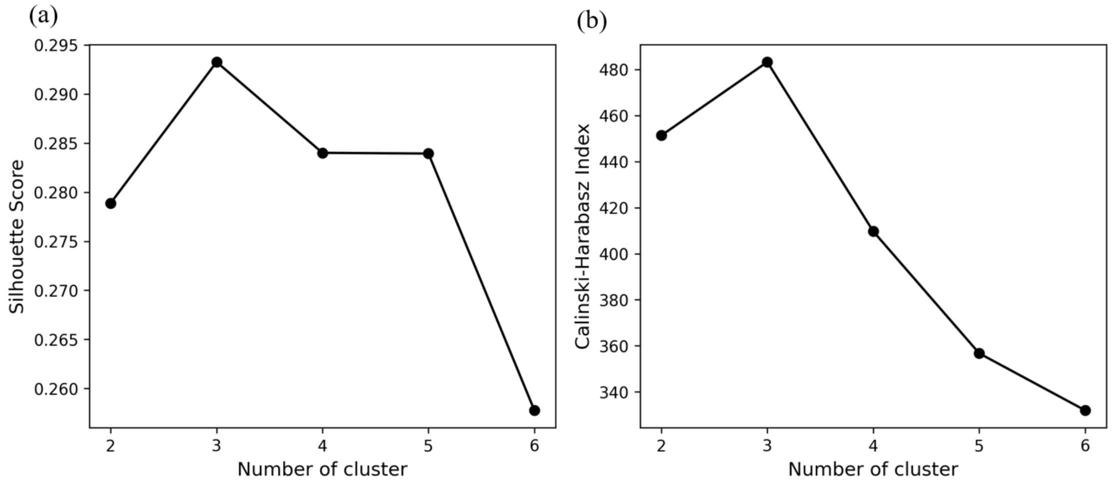


**Figure S1. Validation of the optimal number of clusters.**

Metrics used to determine the optimal number of clusters (K) for the K-means algorithm, evaluated for K ranging from 2 to 6. (a) The silhouette score was plotted against the number of clusters, with the peak score observed at K=3. (b) The Calinski-Harabasz index also showed its highest value at K=3. Based on these two indices, K=3 was selected as the optimal number of clusters.

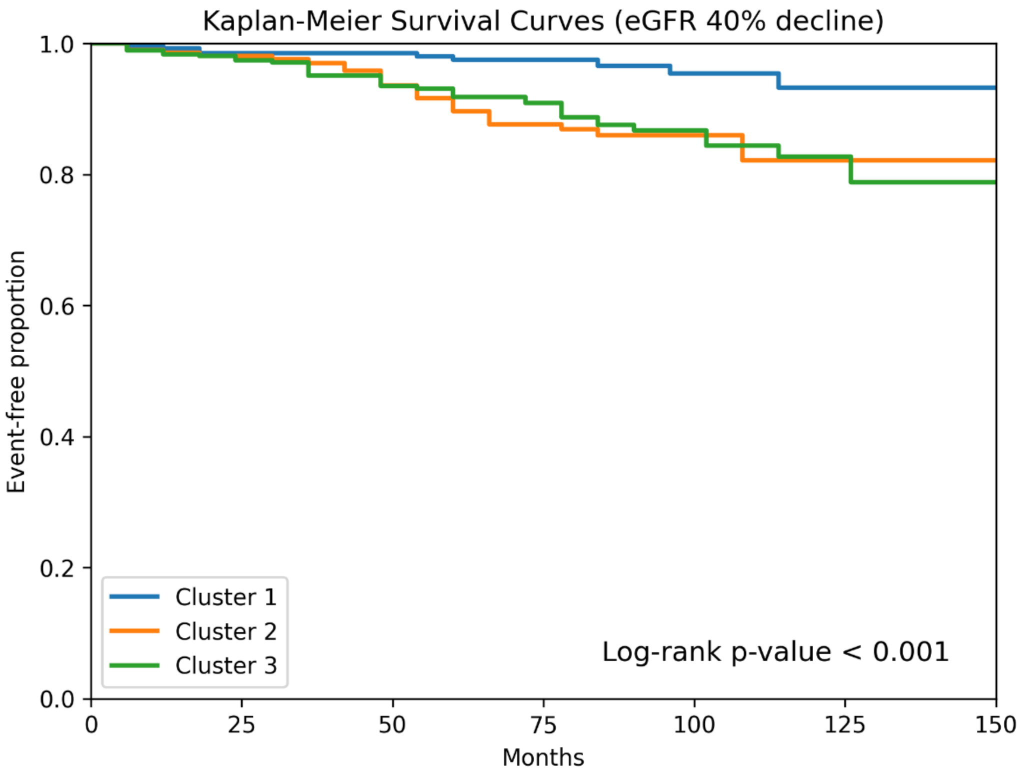


| At risk | | | | | | | |
|---|---|---|---|---|---|---|---|
| Cluster 1 | 284 | 238 | 196 | 131 | 66 | 26 | 3 |
| Cluster 2 | 215 | 186 | 148 | 115 | 53 | 19 | 2 |
| Cluster 3 | 374 | 295 | 232 | 171 | 74 | 21 | 2 |

**Figure S2. Kaplan-Meier curves for a 40% decline in eGFR.**

As a sensitivity analysis, this figure shows Kaplan-Meier curves for the secondary outcome of a ≥ 40% decline in estimated Glomerular Filtration Rate (eGFR) from baseline. The curves compare event-free survival among the three clusters. The p-value from the log-rank test indicates a significant difference among the clusters.

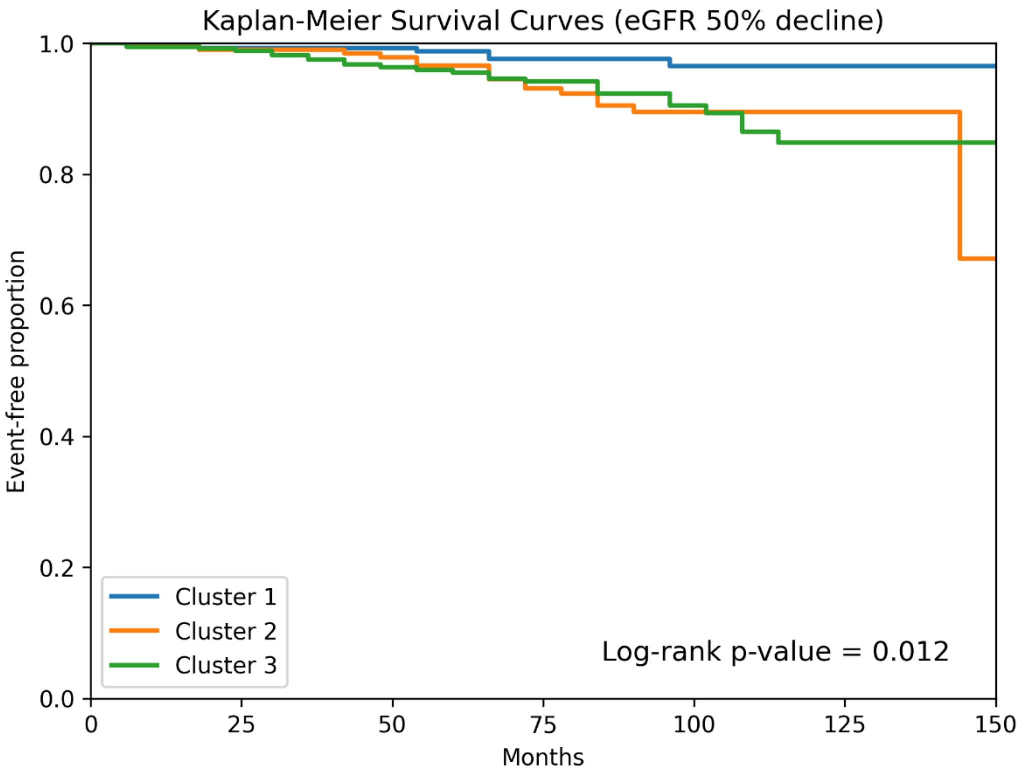


| At risk | | | | | | | |
|---|---|---|---|---|---|---|---|
| Cluster 1 | 284 | 240 | 198 | 132 | 67 | 27 | 3 |
| Cluster 2 | 215 | 187 | 154 | 120 | 54 | 20 | 2 |
| Cluster 3 | 374 | 299 | 238 | 177 | 78 | 22 | 2 |

**Figure S3. Kaplan-Meier curves for a 50% decline in eGFR.**

This figure shows Kaplan-Meier curves for the secondary outcome of a ≥ 50% decline in estimated Glomerular Filtration Rate (eGFR) from baseline, as part of a sensitivity analysis. The event-free survival is compared among the three clusters, and the p-value was determined by the log-rank test.

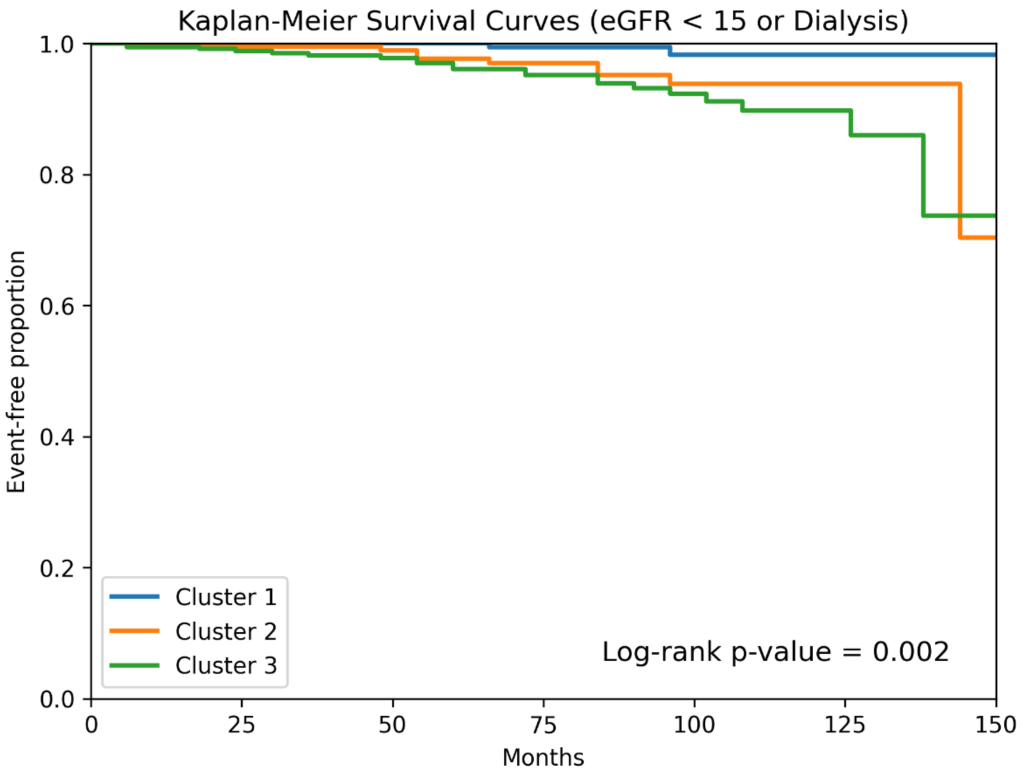


| At risk | 0 | 25 | 50 | 75 | 100 | 125 | 150 |
|---|---|---|---|---|---|---|---|
| Cluster 1 | 284 | 242 | 199 | 135 | 68 | 27 | 3 |
| Cluster 2 | 215 | 188 | 156 | 126 | 57 | 21 | 2 |
| Cluster 3 | 374 | 299 | 242 | 180 | 81 | 24 | 2 |

**Figure S4. Kaplan-Meier curves for kidney failure.**

his figure shows Kaplan-Meier curves for the secondary outcome of kidney failure, defined as a decline in estimated Glomerular Filtration Rate (eGFR) to <15 mL/min/1.73m$^2$ or the initiation of kidney replacement therapy. The event-free survival is compared among the three clusters, with the p-value calculated using the log-rank test.

(a)

Cluster by GMM (columns); Cluster by k-means (rows)

| | 1 | 2 | 3 |
|---|---|---|---|
| **1** | 201 | 44 | 39 |
| **2** | 67 | 148 | 0 |
| **3** | 17 | 47 | 310 |

(b)

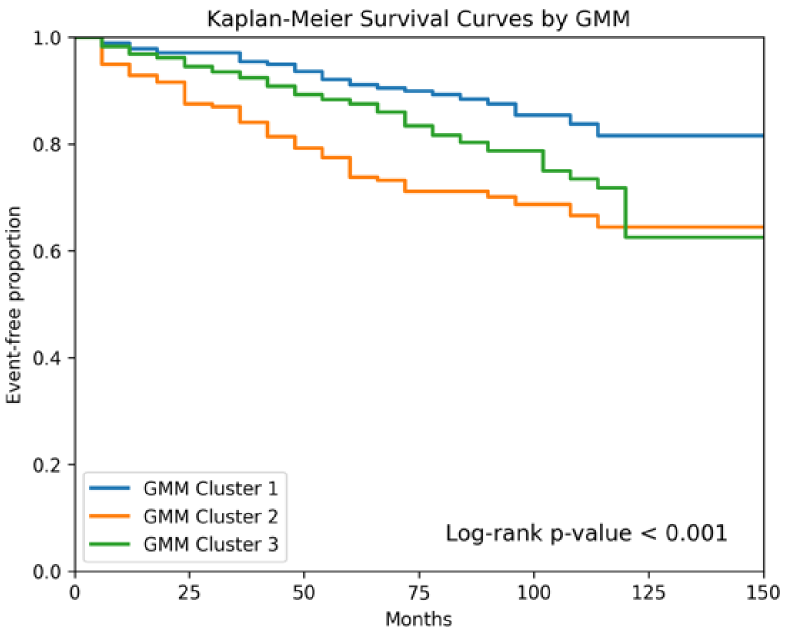


| At risk | | | | | | | |
|---|---|---|---|---|---|---|---|
| GMM Cluster 1 | 285 | 231 | 187 | 130 | 66 | 19 | 2 |
| GMM Cluster 2 | 239 | 184 | 135 | 97 | 42 | 18 | 2 |
| GMM Cluster 3 | 349 | 271 | 210 | 143 | 63 | 21 | 2 |

**Figure S5. Clustering analysis by Gaussian Mixture Model.**

(a) A contingency table showing the overlap in patient classification between the primary K-means clustering and the Gaussian Mixture Model (GMM) clustering. The table shows a high degree of concordance between the two methods, with most patients being classified into corresponding clusters. (b) Kaplan-Meier curves comparing the event-free survival for the primary outcome (≥ 30% eGFR decline) among the three clusters identified by the GMM. The log-rank test demonstrated a significant prognostic separation among the GMM-derived clusters ($p < 0.001$).

**STROBE Statement**—checklist of items that should be included in reports of observational studies

| | Item No | Recommendation | Page No |
|---|---|---|---|
| **Title and abstract** | 1 | (*a*) Indicate the study's design with a commonly used term in the title or the abstract | 2 |
| | | (*b*) Provide in the abstract an informative and balanced summary of what was done and what was found | 2 |
| **Introduction** | | | |
| Background/rationale | 2 | Explain the scientific background and rationale for the investigation being reported | 4-5 |
| Objectives | 3 | State specific objectives, including any prespecified hypotheses | 5 |
| **Methods** | | | |
| Study design | 4 | Present key elements of study design early in the paper | 5-6 |
| Setting | 5 | Describe the setting, locations, and relevant dates, including periods of recruitment, exposure, follow-up, and data collection | 5-6 |
| Participants | 6 | (*a*) *Cohort study*—Give the eligibility criteria, and the sources and methods of selection of participants. Describe methods of follow-up<br>*Case-control study*—Give the eligibility criteria, and the sources and methods of case ascertainment and control selection. Give the rationale for the choice of cases and controls<br>*Cross-sectional study*—Give the eligibility criteria, and the sources and methods of selection of participants | 5-6 |
| | | (*b*) *Cohort study*—For matched studies, give matching criteria and number of exposed and unexposed<br>*Case-control study*—For matched studies, give matching criteria and the number of controls per case | NA |
| Variables | 7 | Clearly define all outcomes, exposures, predictors, potential confounders, and effect modifiers. Give diagnostic criteria, if applicable | 6-7 |
| Data sources/ measurement | 8* | For each variable of interest, give sources of data and details of methods of assessment (measurement). Describe comparability of assessment methods if there is more than one group | 6-7 |
| Bias | 9 | Describe any efforts to address potential sources of bias | 6-7 |
| Study size | 10 | Explain how the study size was arrived at | 6 |
| Quantitative variables | 11 | Explain how quantitative variables were handled in the analyses. If applicable, describe which groupings were chosen and why | 7-9 |
| Statistical methods | 12 | (*a*) Describe all statistical methods, including those used to control for confounding | 8-9 |
| | | (*b*) Describe any methods used to examine subgroups and interactions | NA |
| | | (*c*) Explain how missing data were addressed | 7 |
| | | (*d*) *Cohort study*—If applicable, explain how loss to follow-up was addressed<br>*Case-control study*—If applicable, explain how matching of cases and controls was addressed<br>*Cross-sectional study*—If applicable, describe analytical methods taking account of sampling strategy | 7 |
| | | (*e*) Describe any sensitivity analyses | 9 |

| | | | |
|---|---|---|---|
| **Results** | | | |
| Participants | 13* | (a) Report numbers of individuals at each stage of study—eg numbers potentially eligible, examined for eligibility, confirmed eligible, included in the study, completing follow-up, and analysed | 10, Figure 1 |
| | | (b) Give reasons for non-participation at each stage | 10 |
| | | (c) Consider use of a flow diagram | Figure 1 |
| Descriptive data | 14* | (a) Give characteristics of study participants (eg demographic, clinical, social) and information on exposures and potential confounders | 10, Table 1 |
| | | (b) Indicate number of participants with missing data for each variable of interest | 10 |
| | | (c) *Cohort study*—Summarise follow-up time (eg, average and total amount) | 10 |
| Outcome data | 15* | *Cohort study*—Report numbers of outcome events or summary measures over time | 10 |
| | | *Case-control study*—Report numbers in each exposure category, or summary measures of exposure | |
| | | *Cross-sectional study*—Report numbers of outcome events or summary measures | |
| Main results | 16 | (*a*) Give unadjusted estimates and, if applicable, confounder-adjusted estimates and their precision (eg, 95% confidence interval). Make clear which confounders were adjusted for and why they were included | 12, Table 3 |
| | | (*b*) Report category boundaries when continuous variables were categorized | 7, Figure 3 |
| | | (*c*) If relevant, consider translating estimates of relative risk into absolute risk for a meaningful time period | NA |
| Other analyses | 17 | Report other analyses done—eg analyses of subgroups and interactions, and sensitivity analyses | 12-13, Table 4, Figure S2-5 |
| **Discussion** | | | |
| Key results | 18 | Summarise key results with reference to study objectives | 13-14 |
| Limitations | 19 | Discuss limitations of the study, taking into account sources of potential bias or imprecision. Discuss both direction and magnitude of any potential bias | 15 |
| Interpretation | 20 | Give a cautious overall interpretation of results considering objectives, limitations, multiplicity of analyses, results from similar studies, and other relevant evidence | 15 |
| Generalisability | 21 | Discuss the generalisability (external validity) of the study results | 15 |
| **Other information** | | | |
| Funding | 22 | Give the source of funding and the role of the funders for the present study and, if applicable, for the original study on which the present article is based | 19 |

*Give information separately for cases and controls in case-control studies and, if applicable, for exposed and unexposed groups in cohort and cross-sectional studies.

**Note:** An Explanation and Elaboration article discusses each checklist item and gives methodological background and published examples of transparent reporting. The STROBE checklist is best used in conjunction with this article (freely available on the Web sites of PLoS Medicine at http://www.plosmedicine.org/, Annals of Internal Medicine at http://www.annals.org/, and Epidemiology at http://www.epidem.com/). Information on the STROBE Initiative is available at www.strobe-statement.org.
